# Smart stethoscope for cardiac auscultation in general practice: a prospective feasibility study of AI-assisted detection of atrial fibrillation, heart failure, and valvular heart disease

**DOI:** 10.64898/2026.02.21.26346766

**Authors:** Ralf E. Harskamp

## Abstract

**Objectives:** Artificial intelligence (AI)–enabled digital stethoscopes combine phonocardiography and electrocardiography to support detection of cardiac rhythm and structural abnormalities. This study evaluated the feasibility and exploratory diagnostic performance of AI-guided cardiac auscultation during routine general practice consultations and home visits.

**Methods:** In this prospective feasibility study, 50 consecutive patients aged ≥65 years underwent AI-assisted auscultation using the Eko CORE 500 during routine care. Recordings were attempted at four standard cardiac positions. Feasibility outcomes included technical failure, workflow disruption, and proportion of analyzable recordings (defined as successful AI output based on combined ECG and phonocardiography signals). Exploratory diagnostic performance was assessed against previously established diagnoses of atrial fibrillation (AF), heart failure (HF), or valvular heart disease (VHD) documented in the electronic medical record.

**Results:** AI-guided cardiac auscultation was completed in all patients without device malfunction or meaningful workflow disruption (median acquisition time 1–2 minutes). At least one analyzable recording was obtained in 47/50 patients (94%), and complete four-position analyses in 42/50 (84%). Signal limitations were mainly attributable to obesity, chest hair, or excess breast tissue. Among 47 analyzable patients, 11 had known AF, HF, or VHD. Sensitivity for detecting these conditions was 81.8% and specificity 91.7%. One new case of clinically relevant mitral regurgitation was identified.

**Conclusions:** AI-enabled digital auscultation was feasible in routine general practice, with high rates of analyzable recordings and minimal workflow impact. Larger studies with contemporaneous reference standards are warranted to determine clinical utility.

## INTRODUCTION

Cardiovascular disease remains a leading cause of morbidity and mortality worldwide [1]. While early detection is crucial to enable timely initiation of guideline-directed medical therapy or interventions, most conditions such as valvular heart disease (VHD), atrial fibrillation (AF), or heart failure (HF), are frequently diagnosed at an advanced stage, often after acute deterioration, when opportunities to modify disease progression are limited. [1,2] In many cases, patients first present in general practice with non-specific symptoms (such as dyspnea, fatigue, or palpitations) that do not immediately suggest an underlying cardiac cause. Improving early detection therefore requires solutions that strengthen the GP’s ability to identify possible cardiac disease at the point of care, without disrupting routine workflows. Cardiac auscultation is a core component of physical examination, yet its diagnostic accuracy depends on GP experience and may be limited in busy general practice settings. [3,4] Recent advances in digital signal processing and machine learning have led to “smart stethoscopes” that combine amplified auscultation with electrocardiography (ECG) and automated AI-based analysis [5–7]. These systems aim to assist clinicians by identifying rhythm disturbances and acoustic patterns suggestive of structural heart disease.

Although diagnostic accuracy has been evaluated in controlled or hospital-based settings [8,9], only recently real-world feasibility in community-based healthcare settings have become a topic of interest. This study contributes to this knowledge base, as it involves a prospective feasibility study to assess implementation, analyzability, and exploratory diagnostic performance of AI-guided cardiac auscultation in older adults in routine primary care.

## METHODS

### Study design and setting

This prospective diagnostic feasibility study was conducted between December 2025 and February 2026 at a large primary care center in Amsterdam, the Netherlands. The study followed principles of the STARD-AI reporting guideline [10].

### Participants

Consecutive patients aged ≥65 years attending routine consultations or receiving home visits were eligible. Digital auscultation was incorporated pragmatically into standard care. No additional exclusion criteria were applied.

### Index test

The index test was AI-assisted cardiac auscultation using the Eko CORE 500 digital stethoscope (Eko Health, Emeryville, CA, USA), paired with a smartphone application (FDA-cleared and CE-marked). Recordings were attempted at four standard positions: right upper sternal border, left upper sternal border, left lower sternal border, and apex. The device simultaneously records ECG and phonocardiography signals. Automated AI outputs classify rhythm (e.g., AF detection) and indicate suspected structural abnormalities. When signal quality was insufficient, repeated attempts were made and alcohol wipes to wet the skin were used to optimize acoustic coupling. A recording was considered analyzable if the application generated an AI-based output based on adequate ECG and phonocardiography signals.

### Feasibility outcomes

The primary objective was feasibility during routine care. Feasibility metrics included: completion during consultation or home visit, technical malfunction, clinician-reported workflow disruption, proportion of patients with ≥1 analyzable recording, proportion with complete four-position analyzability.

### Reference standard and diagnostic evaluation

The reference standard was the presence of a previously established diagnosis of AF, HF, or VHD documented in the electronic medical record prior to inclusion. Diagnoses were based on routine clinical assessment, including ECG, imaging, and specialist correspondence when available.

AI results were considered positive if AF or structural heart disease was indicated. Non-analyzable recordings were excluded from diagnostic performance analyses.

Sensitivity, specificity, predictive values, and overall accuracy were calculated descriptively with 95% confidence intervals. No formal sample size calculation was performed, as feasibility was the primary objective.

### Ethics

The Medical Ethics Committee of Amsterdam UMC waived formal review (reference W23_074), as the study involved routine care data collected as part of a quality improvement initiative. The practice setting is an academic healthcare center in which enlisted patients are aware (with an opt out option) that their de-identified data are used for research and quality improvement purposes.

## RESULTS

### Participant characteristics

Fifty patients were included (median age 72 years; 52% female). Eleven patients had previously established AF, HF, or VHD. Patients were for the majority seen in the clinic, and some at the patient’s own home during a home visit. Indications for auscultation included chest discomfort, dyspnea, cough, edema, palpitations, fatigue, but also routine chronic disease follow-up of (very) high-risk patients and/or complex cases. Baseline characteristics are summarized in *Table 1*.

**Table 1.** Baseline characteristics of the study population (n = 50)

| **Characteristic** | **Statistic** |
| --- | --- |
| Age, years, median (IQR) | 72 (68 to 79) |
| Female sex, n (%) | 26 (52.0) |
| Obesity (BMI≥30), n (%) | 16 (32.0) |
| Hypertension, n (%) | 28 (56.0) |
| Diabetes mellitus, n (%) | 11 (22.0) |
| Current or former smoking, n (%) | 13 (26.0) |
| Ischemic heart disease, n (%) | 9 (18.0) |
| Atrial fibrillation, n (%) | 5 (10.0) |
| Heart failure, n (%) | 3 (6.0) |
| Valvular heart disease, n (%) | 3 (6.0) |

### Feasibility

AI-guided auscultation was performed in 50 patients; an example of a recording is shown in *figure 1*. No device malfunctions occurred. Acquisition time was typically 1–2 minutes (maximum 5 minutes), and no meaningful workflow disruption was reported. Obtaining a sufficient ECG signal was most challenging. Applying (more than normal) pressure on the stethoscope for good skin contact as well as steady hand positioning was essential to obtain sufficient quality ECG recordings. Moreover, patients who had just applied creams and skin lotions appeared to interfere with the ECG signal, thus sometimes alcohol wipes were needed to improve signal quality. In the end, at least one analyzable recording was obtained in 47/50 patients (94%). Complete four-position analyzability was achieved in 42/50 patients (84%). In three patients, no analyzable recordings were obtained despite repeated attempts. Non-analyzable positions were most frequent at the left lower sternal border and apex. Observed interfering factors included obesity, chest hair interfering with ECG contact, and excess breast tissue affecting apical recordings.

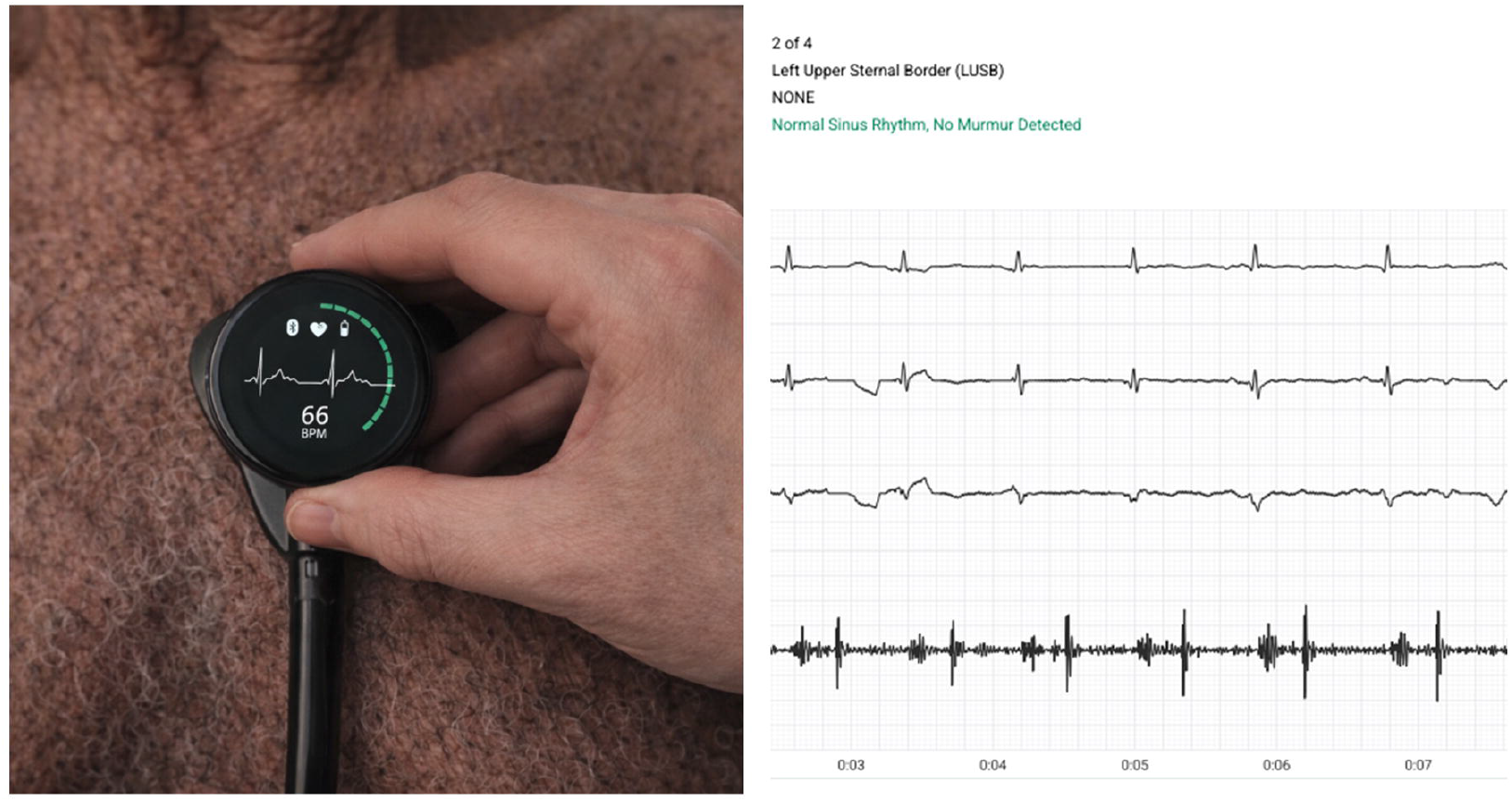

### Exploratory diagnostic performance

Among 47 patients with analyzable recordings, 11 had a known diagnosis of AF, HF, or VHD. AI outputs were concordant in 9 of these 11 cases. Two false negatives occurred: one patient with paroxysmal AF not in AF during recording, and one patient with HF with preserved ejection fraction. Three false positives were observed: one clinically insignificant murmur, two AF alerts attributed to rhythm irregularity and baseline noise. Sensitivity was 81.8%, specificity 91.7%, with high negative predictive value (Table 2). In addition, AI-assisted auscultation prompted further evaluation in one patient in his late 70s presenting with fatigue. Subsequent echocardiography confirmed moderate-to-severe mitral regurgitation.

**Table 2.** Diagnostic properties for detecting previously established cardiac conditions (n = 47)

| **Statistic** | **Value (95% CI)** |
| --- | --- |
| Sensitivity | 81.8% (48.2-97.7) |
| Specificity | 91.7% (77.5-98.3) |
| Positive likelihood ratio | 9.8 (3.2-30.1) |
| Negative likelihood ratio | 0.2 (0.1-0.7) |
| Positive predictive value | 75.0% (49.5-90.2) |
| Negative predictive value | 94.3% (82.4-98.3) |
| Accuracy | 89.4% (76.9-96.5) |

True positives, n=9; false positives, n=3; false negatives, n=2; true negatives, n=33.

## DISCUSSION

In this prospective feasibility study, AI-enabled digital auscultation was readily integrated into routine general practice consultations and home visits. Analyzable recordings were obtained in the vast majority of older patients, and the procedure required minimal additional time.

The findings align with prior studies demonstrating promising diagnostic performance of AI-based auscultation in controlled settings [11–13]. However, implementation in primary care poses distinct challenges, including time constraints, signal acquisition in diverse patient populations, and integration into existing workflows [3,4].

Signal acquisition was feasible in most cases, though anatomical factors such as obesity and breast tissue limited analyzability at certain positions. These predictable barriers are relevant for real-world implementation.

Large pragmatic trials provide important context. In the TRICORDER cluster-randomized trial, broad implementation of an AI stethoscope did not significantly increase HF detection overall, but per-protocol analyses showed higher detection rates and shorter time to diagnosis when the device was actually used [14]. These findings suggest that clinical impact depends not only on algorithm performance, but also on targeted use, workflow integration, and clinician engagement [15].

This study has limitations. It was conducted at a single site with a modest sample size and used previously established diagnoses as reference standards rather than systematic contemporaneous testing. The performing GP was not blinded to medical history, reflecting intended real-world use but introducing potential bias.

Nevertheless, this study provides early evidence that AI-assisted auscultation is operationally feasible in routine primary care and home-based settings among older adults.

## CONCLUSIONS

AI-enabled smart stethoscope–guided auscultation was feasible in routine primary care, with high rates of analyzable ECG and phonocardiography recordings and minimal workflow disruption. Exploratory diagnostic performance was encouraging. Larger studies incorporating systematic reference standards and structured implementation strategies are needed to define clinical effectiveness and impact on patient outcomes.

## Data Availability

All data produced in the present work are contained in the manuscript

